# Implementation of One Health surveillance systems: opportunities and challenges - Lessons learned from the OH-EpiCap application

**DOI:** 10.1101/2023.11.02.23297972

**Authors:** Henok Ayalew Tegegne, Frederick T. A. Freeth, Carlijn Bogaardt, Emma Taylor, Johana Reinhardt, Lucie Collineau, Joaquin M Prada, Viviane Hénaux

## Abstract

As the complexity of health systems has increased over time, there is an urgent need for developing multi-sectoral and multi-disciplinary collaboration within the domain of One Health (OH). Despite the efforts to promote collaboration in health surveillance and overcome professional silos, implementing OH surveillance systems in practice remains challenging for multiple reasons. In this study, we describe the lessons learned from the evaluation of OH surveillance using OH-EpiCap (an online evaluation tool for One Health epidemiological surveillance capacities and capabilities), the challenges identified with the implementation of OH surveillance, and the main barriers that contribute to its sub-optimal functioning, as well as possible solutions to address them. We conducted eleven case studies targeting the multi-sectoral surveillance systems for antimicrobial resistance in Portugal and France, *Salmonella* in France, Germany, and the Netherlands, *Listeria* in The Netherlands, Finland and Norway, *Campylobacter* in Norway and Sweden, and psittacosis in Denmark. These evaluations facilitated the identification of common strengths and weaknesses, focusing on the organization and functioning of existing collaborations and their impacts on the surveillance system. Lack of operational and shared leadership, adherence to FAIR data principles, sharing of techniques, and harmonized indicators led to poor organization and sub-optimal functioning of OH surveillance systems. In most cases, the effectiveness of OH surveillance over traditional surveillance, operational costs, behavioural changes, and population health outcomes brought by the OH surveillance have not been evaluated. To this end, the establishment of a formal governance body with representatives from each sector could assist in overcoming long-standing barriers. Moreover, demonstrating the impacts of OH-ness of surveillance may facilitate the implementation of OH surveillance systems.

## Introduction

Recent disease emergences (Ebola, Zika, avian influenza, Covid-19) have reinforced global attention to the importance of integrated infectious disease surveillance systems and the application of the One Health (OH) approach at all levels [1-3]. The approach mobilizes multiple sectors and disciplines, and ensures communication, collaboration, and coordination among all relevant ministries, agencies, stakeholders, sectors, and disciplines, for optimal action [4]. OH surveillance is a collaborative and systematic collection, validation, analysis, interpretation of data, and dissemination of information collected on humans, animals, and the environment to inform decisions for more effective health interventions [5, 6]. In practice, different countries have implemented the principle of OH surveillance with varying successes and challenges [7].

While there has been a wide-ranging commitment to the OH approach for addressing complex health problems by several national and international organizations and professional bodies, its operationalization has so far proved to be challenging [8]. Implementation is often a complex issue requiring collaboration between diverse and multi-disciplinary partnerships [1] and mostly occurs in crisis times [8]. For instance, most countries lack formal mechanisms for the coordination and integration of activities across the human health, agricultural, and environmental sectors, which are traditionally based in separate ministries or government agencies with different mandates on activities and spending. As a result, practical applications of OH approaches have largely been ad-hoc [8, 9].

There is a need for regular evaluation of the performance of OH surveillance to identify the main challenges to the effective implementation of OH surveillance [10, 11]. In this regard, we developed a generic OH surveillance evaluation tool, OH-EpiCap [12], to characterize and assess epidemiological surveillance capacities and capabilities in existing national surveillance systems. This tool is a standalone, user-friendly, web application developed to conduct an evaluation of multiple aspects related to the organization of the OH surveillance system, its functioning, and its impacts on the surveillance and beyond [12-14]. It supports the diagnosis of strengths and weaknesses in multi-sectoral collaborations and helps to improve collaborative activities at all steps of surveillance [15]. In this study, we assessed the OH practices of foodborne and other zoonotic hazards surveillance systems in several European countries to identify the main barriers that contribute to sub-optimal OH functioning.

## Methodology

### Study design

Eleven national multi-sectoral surveillance systems were evaluated using the OH-EpiCap tool between April and November 2022. These surveillance systems focus on foodborne and other emerging zoonotic hazards in European countries: antimicrobial resistance (AMR) in France and Portugal, *Salmonella* in Germany, France and The Netherlands, *Listeria* in Finland, Norway and The Netherlands, *Campylobacter* in Norway and Sweden, and psittacosis in Denmark. These case studies were identified in the framework of the OH European Joint Programme MATRIX project.

The OH-EpiCap tool [12] is a semi-quantitative evaluation tool, organized around three dimensions, each of them divided into four targets, Figure 1. The first dimension relates to the organization of the OH surveillance system, including the formalization of the system, coverage, availability of resources, and evaluation. The second dimension assesses operational activities such as data collection and methods sharing, data sharing, data analysis and interpretation, and communication. The last dimension evaluates the impacts of the OH surveillance system and comprises targets for technical outputs, collaborative added-value, immediate and intermediate outcomes, and ultimate outcomes. Each target score is calculated from four indicators. A standardized scoring guide details, for each indicator, the possible scores and how they should be awarded [16].

**Figure 1:**
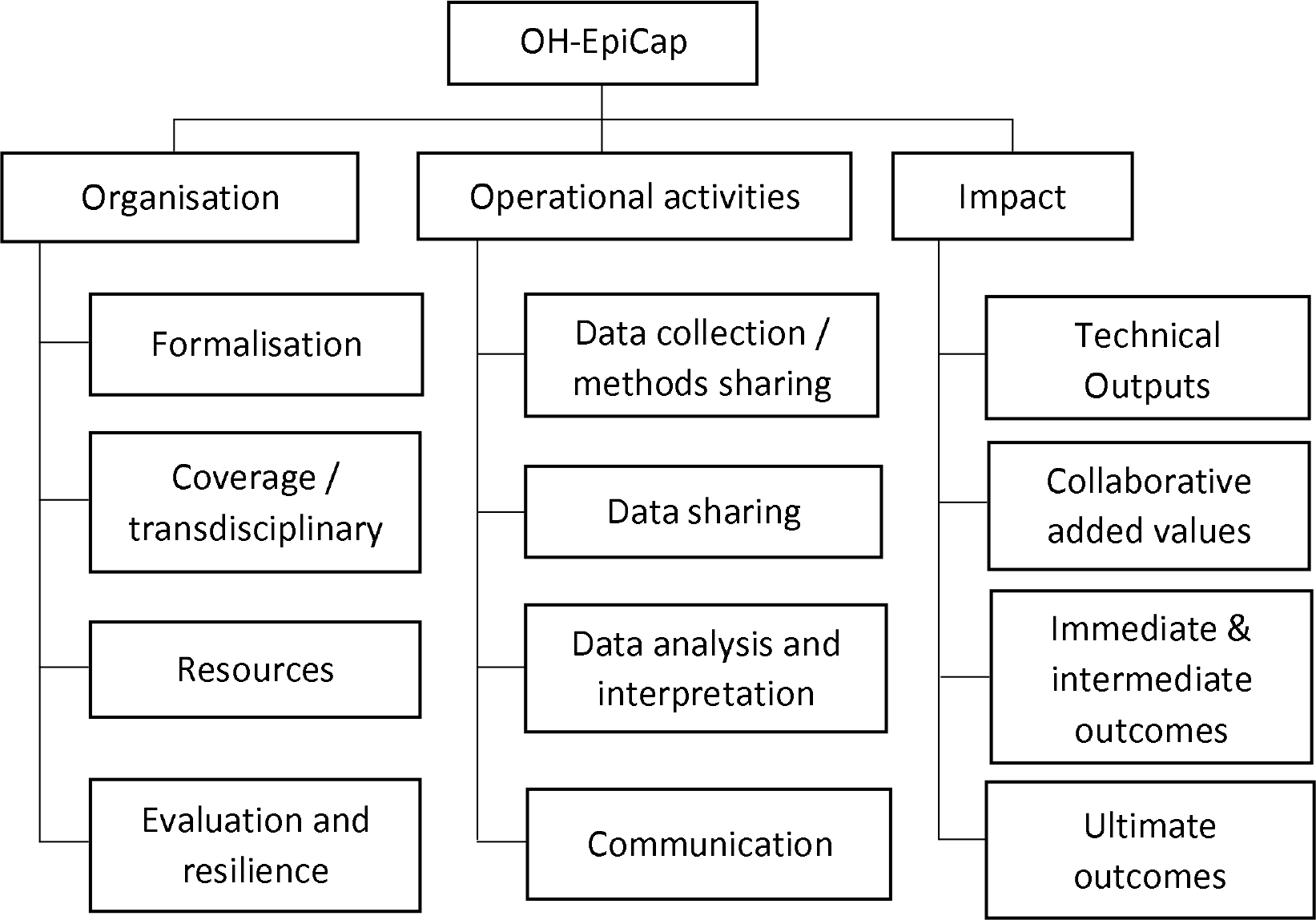
Structural overview of the OH-EpiCap targets, grouped by dimension.

OH-EpiCap evaluation of a surveillance system takes place through a half-day workshop gathering a panel of surveillance representatives from the sectors and disciplines relevant to the hazard of interest. For each question in the questionnaire, the panel jointly agrees on the answer that best represents their system between four options, which is then transformed into a semi-quantitative scale ranging from 1 to 4 (higher values suggesting better adherence to OH principles (better integration of sectors), and lower values suggesting improvements may be beneficial). The panel answers each question using the Shiny application (https://freddietafreeth.shinyapps.io/OH-EpiCap/), which summarizes and visualizes the results in the form of a report with interactive figures. If a workshop cannot be organised, the questionnaire can be filled sequentially by the surveillance representatives from each sector, with a back-and-forth process to reach a consensus.

Representatives of the surveillance systems of psittacosis in Denmark, *Listeria* in Finland, *Salmonella* in Germany, and *Campylobacter* in Sweden carried out the evaluation through a half-day workshop. Other evaluations were conducted through the completion of the questionnaire, either sequentially by experts from several sectors of surveillance (e.g. AMR in Portugal, AMR in France, *Campylobacter* in Norway), or by one to two experts but who had a good knowledge of the organization of the surveillance and collaborations across sectors (e.g. *Salmonella* in France, *Listeria* and *Salmonella* in the Netherlands, and *Listeria* in Norway). A list of the institutes that participated in each OH-EpiCap evaluation is provided in supplementary file 1. The Results and Discussion section below presents the main weaknesses and strengths identified from the OH-EpiCap scores and the contextual and interpretive comments provided by the evaluators (surveillance representatives).

## Results and Discussion

Our study focused on existing collaborative practices in eleven multi-sectoral surveillance systems targeting foodborne and emerging zoonotic hazards in various European countries. Overall OH-EpiCap scores across the eleven systems evaluated were between 32.4 and 86.1 out of 100.0, with a mean score of 49.0 and a standard deviation of 17.7, indicating a wide spread of results, Figure 2. Systems scoring high overall generally had high scores in Dimension 1 (Organization). Systems with low scores scored roughly the same across all three dimensions. Several of the highest-scoring systems’ weakest dimension was Dimension 2 (Operational activities); however, System 9 did not follow this trend and scored roughly equally across Dimensions 1 and 3, with Dimension 2 being higher. Middle-scoring systems commonly exhibited weaknesses in Dimension 3 (Impact), indicating this area requiring improvement.

**Figure 2:**
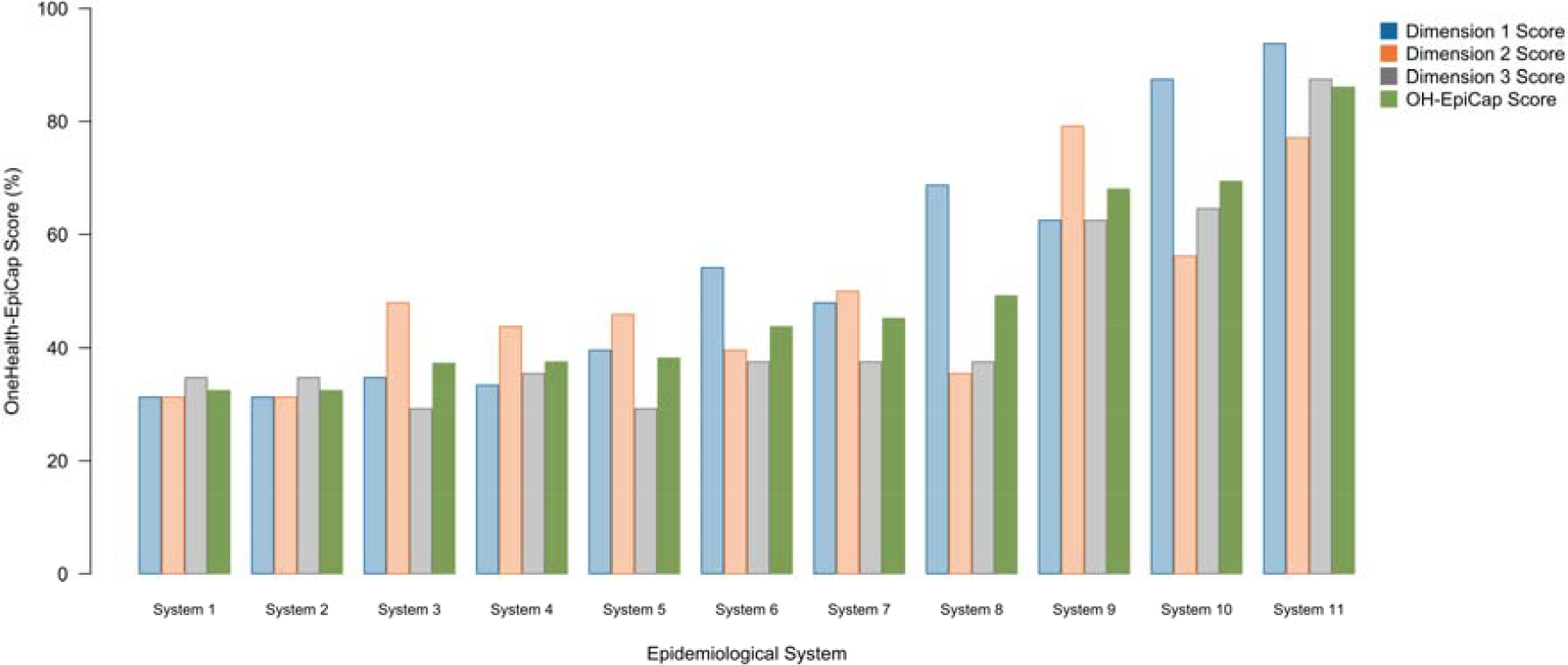
Dimension-specific (Dimension 1: Organization – blue, dimension 2: Operational activities – orange, and dimension 3: Impact – grey) and overall (green) OH-EpiCap scores for each epidemiological system evaluated.

Overall, the median score of the targets across the systems evaluated were between 2 and 3, with the exception of the target *Coverage/Transdisciplinary*, which exceeded 3, Figure 3. Moreover, the lower and upper quartiles for the *Coverage/Transdisciplinary* target were also the highest, ranging from 2.5 to 3.5. The *Communication* target also scored consistently high across all systems, indicated by a narrow interquartile range just under 3. The four targets scoring low on average were *Formalization, Evaluation and Resilience, Technical Outputs*, and *Ultimate Outcomes*, each with a median score of 2. OH-EpiCap dimensions visualised at the indicator level are provided in supplementary file 2.

**Figure 3:**
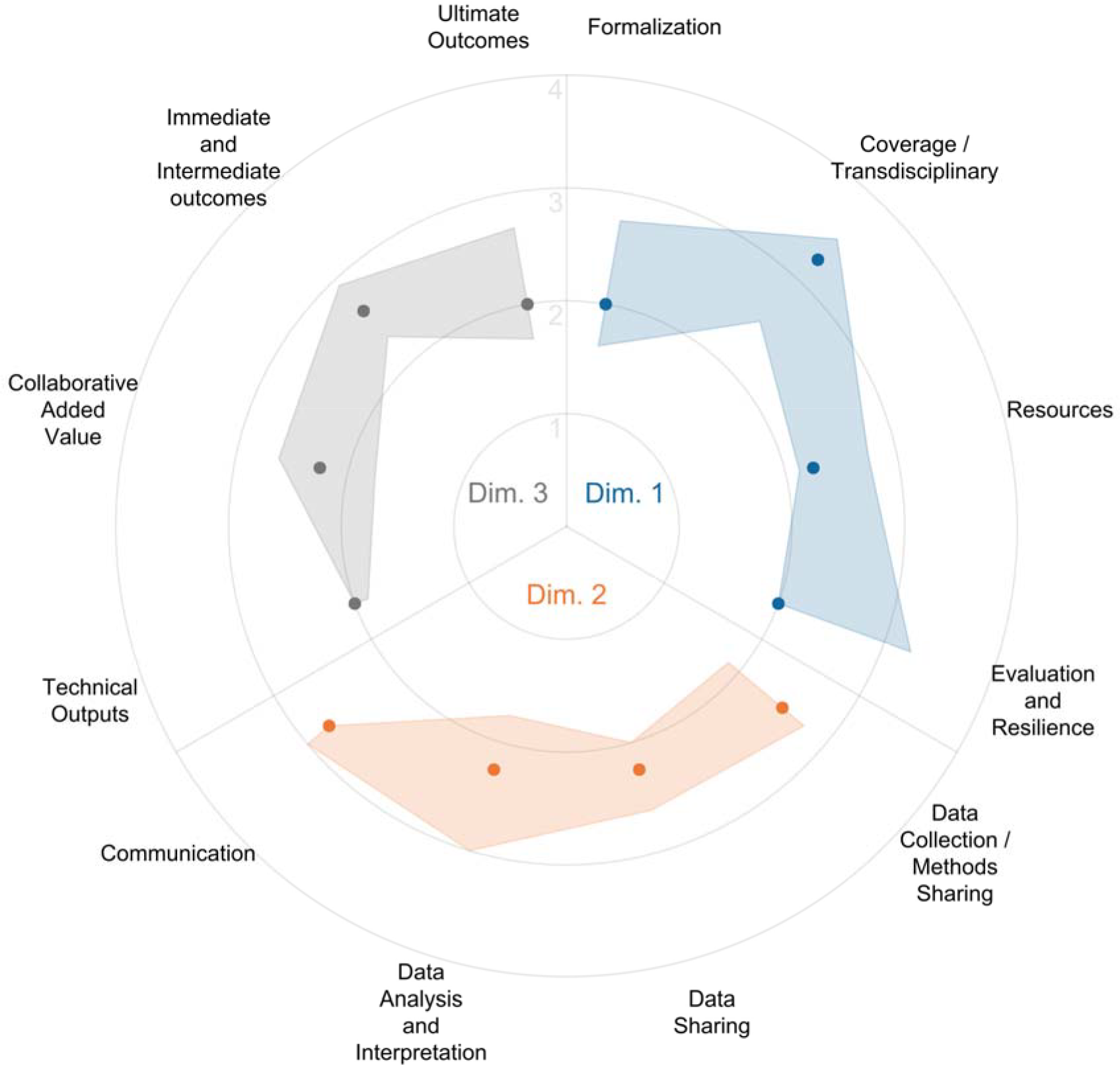
OH-EpiCap dimensions visualised at the target level. Plotted points indicate the median score with interquartile range indicated by the shaded polygons across all systems investigated. Dim1 – Dimension 1: Organization, Dim2 – Dimension 2: Operational activities and Dim3 – Dimension 3: Impact.

### Dimension One - Organization

Regarding *Formalization*, some evaluated surveillance systems established a cross-sectoral aim (median [interquartile range]: 3 [1.5-3]) through participative approaches or from inter-ministerial documents, which may not, however, fully meet all stakeholder expectations. Various types of supporting documentation (2 [2-2]) was reported, including legislation, strategy papers, and inter-ministerial commitments. However, these documents often cover only specific aspects of OH surveillance. Our findings revealed that supporting documentation is typically shared across sectors and institutions. However, it seems to be insufficient for the proper formalization of OH collaborations. Legislative support is critical for OH formalization, as it provides guidelines for coordinating and integrating surveillance programs across sectors [8, 9]. Additionally, there is a lack of operational and shared leadership (1 [1-3]). Previous studies emphasized the importance of establishing a steering committee to manage OH activities and engage decision-makers, ministers, and officials from all sectors [2, 3]. Bridging human, animal and environmental health requires multidisciplinary and cross-sectoral leadership. This leadership must address collaboration gaps, minimize duplication, and prevent divisions and isolation [17, 18]. A set of recommendations regarding OH governance, considering important elements such as transparency, accountability, and responsibility, was proposed by experts from the OH-EJP COHESIVE project in their guidelines for establishing a OH risk analysis system for zoonoses [19].

In terms of *Coverage and transdisciplinarity*, the evaluated surveillance systems encompassed relevant sectors (3 [3-4]) and disciplines (3 [3-3]), yet often neglected the environmental sector. Although OH should rely on the triad of human, animal, and environmental health, the latter is often overlooked [20-22]. More specifically, the wildlife component and related ecological issues such as community ecology, evolutionary ecophysiology, and environmental science (soil and climate) were neglected [23, 24]. In recent years, several efforts have been made to engage the environmental sector for collaborative OH action. Thus, in 2022, the United Nations Environment Programme (UNEP) joined the tripartite collaboration as an equal partner to implement OH approach [25]. Moreover, our study also identified a limited representation of economics and social science in the evaluated surveillance systems. Similarly, previous research has highlighted the marginalization of social, legal, and economic sciences within OH [23, 26]. Yet, economics and social sciences have recently gained recognition as essential pillars for developing sustainable strategies against zoonotic diseases [10, 26].

Coverage of actors was more variable (3 [2.5-4]). Implementation of OH surveillance requires integration of various public, private, political, civil society, and business sectors [2]. Identifying and early engaging all actors involved in surveillance can help build trust and create a conducive environment for acceptable and sustainable collaborative solutions [27, 28]. Mapping of the surveillance programs and stakeholders is therefore recommended to identify their roles and missions and characterize the interactions between them [27-30]. Geographical coverage and inclusion of less represented populations varied between evaluated systems (3 [2-4]). Yet, the reinforcement of links with surveillance systems focusing on similar hazards (e.g. mosquito-borne diseases, abortive cattle diseases) has been shown to increase surveillance capacities through the sharing and joint analysis of data, favouring preparedness and rapid mitigation response [31].

Regarding *Resources*, our results showed the existence of OH-focused training opportunities (2 [1.5-3.5]), although they remain insufficient and not specific to the surveillance of the hazard of interest. Organizing joint communication training or pre-emptive collaborative training (such as outbreak simulation exercises) [32] and reinforcing OH education are important to foster multi-sectoral collaborations [3, 19, 33]. OH training should also include cross-cultural communication skills, team building and trust development [18].

Regarding *Evaluation*, internal evaluations are rarely carried out or only partially (2 [1-3]). Similarly, only some systems conducted external evaluations (2 [1-3]) through national authorities or international health agencies like EFSA and ECDC. Some evaluation-based corrective measures were implemented (2.5 [2-3]), underlying the interest of those evaluations. The OH-EpiCap evaluation framework [12] provides a standalone tool for surveillance representatives to conduct an internal evaluation in a short time (half-day workshop). A more thorough evaluation of the weaker OH aspects can then be considered using evaluation tools dedicated to the functioning and performance of surveillance [34] and/or OH aspects [35]. Moreover, the OH-LabCap tool (https://onehealthejp.eu/jip-ohharmony-cap/) supports the assessment of OH capacities related to laboratory activities.

### Dimension Two - Operational activities

Regarding *Data collection and methods sharing*, some joint data warehouses between sectors were set up (2 [2-3]), in particular for sequencing or notification data. These are accessible to relevant stakeholders to upload or download data, or only to access data. Examples of joint aggregated surveillance databases are presented in the literature [42-44].

Regarding *Data sharing*, our study revealed a lack of adherence to FAIR data principles (2 [1-2.5]). Despite the recognized importance of FAIR data, practical application remains challenging [36-38]. Numerous obstacles, including ethical, organizational, legal, technical, political, and economic issues, hindered timely and effective data sharing between health authorities and stakeholders [2, 19, 38, 39]. Common best practices for data collection, sharing, analysis, interpretation, and dissemination were reviewed in recent studies [19, 29]. Additionally, our study found that systematic data quality evaluation (2 [2-3]) was not systematically conducted. EFSA underlined the importance of enhancing data quality and interoperability to better leverage existing data for risk assessment and preparedness within the OH framework [40]. In the evaluated systems, data quality evaluation approaches included proficiency tests, ISO/IEC 17025 accreditation, or dedicated tools. These approaches facilitate data monitoring and communication with providers and stakeholders [41].

Regarding *Data analysis and interpretation*, our study showed limited technique sharing (1.5 [1-2]), and the use of few or no harmonized indicators (2 [1-3]) among sectors. Surveillance representatives reported joint analysis of data from multiple sectors or joint interpretation of results from several sector-specific analyses (2 [2-3]), mostly during outbreak investigations or in collaborative projects. Accordingly, such joint analyses of data favour preparedness and rapid mitigation response [31]. Moreover, combining data from different syndromic surveillance components showed promise for improving the surveillance of foodborne disease outbreaks in humans [45].

In terms of *Communication*, most of the evaluated systems reported that internal communication across sectors (3 [2-3]) could be improved, in particular through the implementation of a formal system. Inter-disciplinary and OH glossaries help overcome communication hurdles (due to sector-specific terminology) and harmonize information exchange between sectors [19, 46, 47]. Joint external communication appears well developed (3 [3-3.5]), while information dissemination to decision-makers occurs mostly within sector (2 [2-3]). The development of operational dashboards can facilitate the sharing of raw data and sensitive information internally and to decision makers through password restricted access, and the display of aggregated and digested information to the large public, using an open access [48].

### Dimension Three - Impact

Regarding *Technical Outputs*, our evaluations revealed that, in most cases, the effectiveness (1 [1-1]) of OH surveillance (over traditional approaches) and its operational costs have not been formally assessed using comprehensive methods (1 [1-1]). While OH surveillance has been in place for over a decade with the expectation of improving efficiency by reducing overlaps among sectors, systematic evaluation of key attributes remains limited [18, 39, 49]. Surveillance representatives indicated that multi-sectoral preparedness and response capacity are in place (3 [3-4]) but emphasized the need for faster and more sensitive responses. Although large outbreaks are often detected in real-time, retrospective (WGS) analyses revealed undetected emergencies in some sectors. Overall, we found that the OH approach has enhanced surveillance effectiveness by detecting more outbreak sources and providing insights into the genetic diversity and clustering of foodborne hazard isolates from different sectors, particularly through WGS data sharing. It has also improved the knowledge of the epidemiological situation (3 [3-3.5]). However, the costs of OH surveillance were found to be increased due to integrated molecular surveillance, additional personnel time with the arrival of WGS analysis, and epidemiological investigations in case of links between human cases and production facilities. Despite being resource-intensive, joint surveillance efforts have also shown economic benefits [39]. Some studies proposed outcome metrics and methodological frameworks for evaluating OH interventions [49-51].

Regarding *Collaborative added value*, although official OH teams (3 [1.5-3]) or networks (2 [1-3.5]) are not well established, collaboration and trust among different sectors have grown through OH surveillance activities. Challenges to foster multidisciplinary collaboration and build operational OH networks are detailed by Khan et al. [21]. We found that international collaborations for hazard surveillance remain sector-dependent (2 [2-3]), relying on specific networks and international health agencies: EFSA and ECDC for the animal and human sectors, and the European Environment Agency (EEA), European Chemicals Agency (ECHA), and EU Commission for the environmental sector. For AMR, effective international collaboration occurs through the Joint Inter-Agency Antimicrobial Consumption and Resistance Analysis (JIACRA), involving the human and animal sectors [40] and global action plans encompassing human, animal, and environmental sectors [52]. Common strategic plans for other hazards are in progress (1 [1-4]), with guidance provided by the quadripartite OH Joint Plan of Action [25] and the Generalizable OH Framework, which compiles resources for OH development from local to international levels [28].

Regarding *Immediate, intermediate and ultimate outcomes*, advocacy efforts were implemented to influence public policy, laws and to educate target groups (government officials, students in health education, and the public) about the mitigation of hazard risk based on information from the OH surveillance (3 [2.25-3.75]). However, its contribution to increasing awareness and understanding of hazards remained limited for some stakeholders (2 [2-3]). Conducting Knowledge, Attitude, and Practices surveys can help identify this contribution [53]. OH surveillance led to some policy changes (2.5 [1.25-3]), such as updating shelf life for risk products and revising antimicrobial use policies in veterinary medicine and humans. Research programs based on multi-institutional consortia, involving multiple disciplines, enable the application of holistic and integrated approaches to tackle the complexity of questions at the interface between human, animal and environmental health.

### OH-EpiCap limits

Although this sample of surveillance systems is not representative of the diversity of situations in OH surveillance, it provides a large overview of strengths and weaknesses regarding multiple aspects (organisation, operational activities, and impact) of OH. The power of OH-EpiCap evaluations relies on the free sharing and exchange of points of view and perceptions by representatives from all sectors and disciplines involved in the surveillance. Accordingly, the panels of surveillance representatives who conducted the evaluations were selected to cover all relevant sectors for the hazard under surveillance. Yet, including the environmental sector was not always feasible especially when this sector was not or had little involvement in the integrated system. Additionally, the surveillance representatives had different expectations and various levels of knowledge about the surveillance system. However, biases and subjectivity in the scoring is limited by the use of a standardized scoring scale and by the fact that the panel needs to reach consensus to answer each indicator.

## Conclusions

Our study evaluated various integrated surveillance systems across Europe, offering a comprehensive overview of strengths and areas for improvement in OH functioning. While European laws and standards influence national policies, each country tailors its surveillance and OH regulations to its unique epidemiological situation and ongoing context (technical infrastructure, surveillance capacity, policy support, etc.). Each surveillance system is inherently distinct, and its potential for evolution towards more integration will depend on the expectations of the various actors and their willingness and capacity to further develop and enhance collaborations at all steps of the surveillance. The OH-EpiCap tool is generic and standalone and we therefore encourage surveillance representatives to conduct such an evaluation of their system to identify their best way forward. Furthermore, given the simplicity and limited time required, we recommend conducting such OH-EpiCap evaluations regularly to monitor the evolution of OH practices over time and their impact on health at the human-animal-environment interface.

## Data Availability

All data produced in the present study are available upon reasonable request to the authors

## Author contributions

VH and JP conceptualized the study. HT, JR, LC, and VH conducted the case studies. CB, FF, ET, and JP developed and maintained the web application for data collection and visualization. HT and VH drafted the manuscript, and all authors participated in editing, reviewing, and approving the final version.

## Confidentiality and ethical statement

The MATRIX project obtained ethical approval from the advisors of the One Health European Joint Programme. Participants were informed via email and verbally about: 1) The voluntary nature of using the OH-EpiCap tool and application; 2) Compliance of the OH-EpiCap tool with the European General Data Protection Regulation, ensuring no collection of personal information; 3) Non-retention of data related to the evaluated OH surveillance system by the web application.

## Declarations of interest

None

## Conflict of interest

The authors declare that the research was conducted in the absence of any commercial or financial relationships that could be construed as a potential conflict of interest.

## Funding

This work was supported by funding from the European Union’s Horizon 2020 Research and Innovation programme under grant agreement No 773830: One Health European Joint Programme. Funding sources did not affect the design of this study, data collection, data analysis, decisions on publication, or preparation of the manuscript.

## Data Availability

The R code and anonymised data used to generate the figures in this paper, and the supplementary figures, can be found in the following GitHub repository https://github.com/FreddieTAFreeth/OH-EpiCap-Cross-System-Analysis.

## Acknowledgments

We thank the surveillance representatives who participated in the OH-EpiCap evaluations.

## Supplementary files

**Supplementary table 1:** List of the institutes that participated in the OH-EpiCap evaluation of their specific surveillance system

**Supplementary file S2:** OH-EpiCap dimensions visualised at the indicator level. Plotted points indicate the median score with interquartile range indicated by the shaded polygons across four targets

## Notes

### Competing Interest Statement

The authors have declared no competing interest.

### Funding Statement

This work was supported by funding from Horizon 2020 Research and Innovation programme under grant agreement No 773830: One Health European Joint Programme. Funding sources did not affect the design of this study, data collection, data analysis, decisions on publication, or preparation of the manuscript.

### Author Declarations

The MATRIX project obtained ethical approval from the advisors of the One Health European Joint Programme.

