## Supplementary File S2 for "Implementation of One Health surveillance systems: opportunities and challenges - Lessons learned from the OH-EpiCap application"

**Supplementary file S2:** OH-EpiCap dimensions visualised at the indicator level. Plotted points indicate the median score with interquartile range indicated by the shaded polygons across four targets

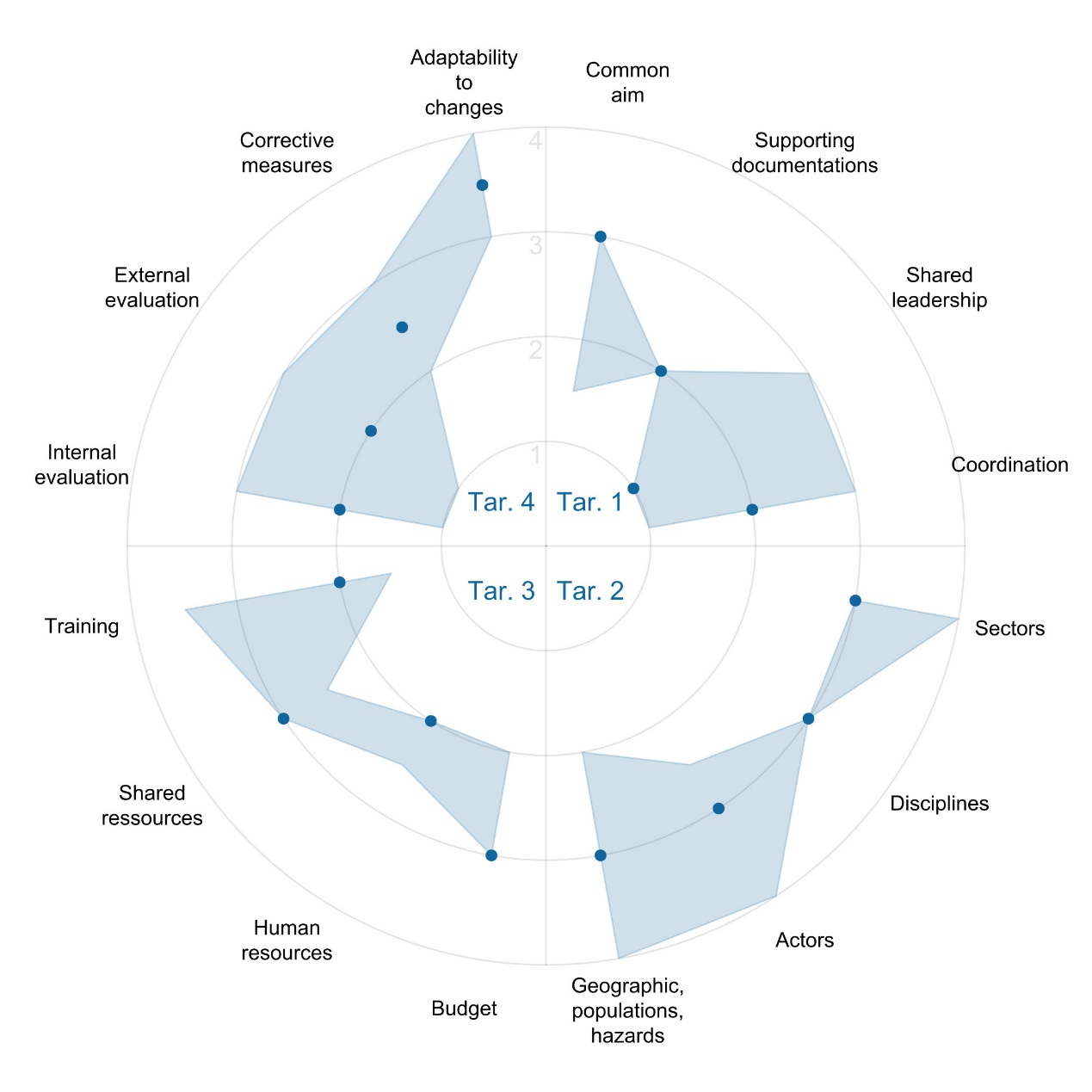

Figure S2_1: OH-EpiCap Dimension one (organization) visualised at the indicator level. Plotted points indicate the median score with interquartile range indicated by the shaded polygons across four targets: target 1: formalization; target 2: coverage and transdisciplinary; target 3: availability of resources; target 4: evaluation.

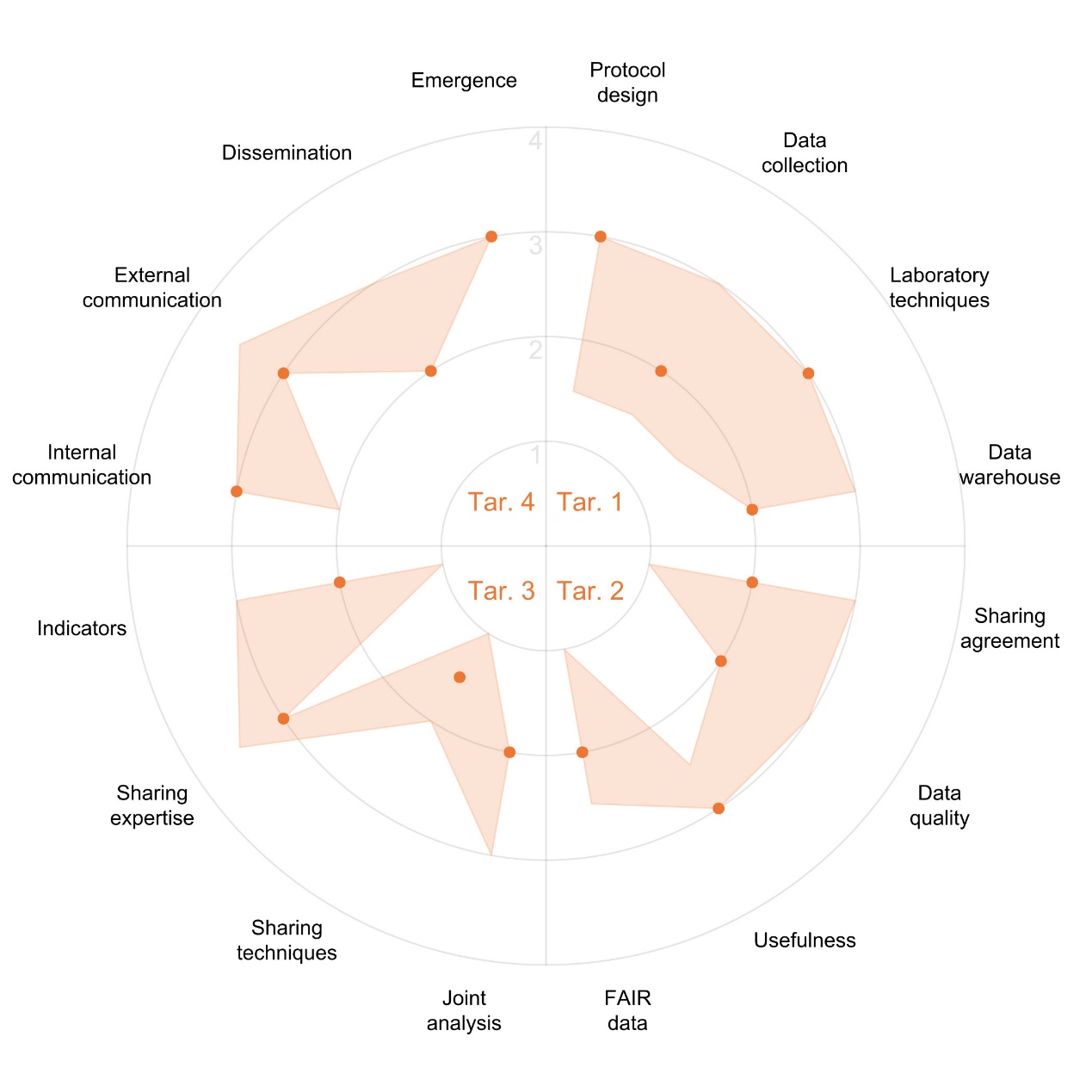

Figure S2_2: OH-EpiCap Dimension two (operation) visualised at the indicator level. Plotted points indicate the median score with interquartile range indicated by the shaded polygons across four targets: target 1: data collection and methods sharing; target 2: data sharing; target 3: data analysis and interpretation; target 4: communication.

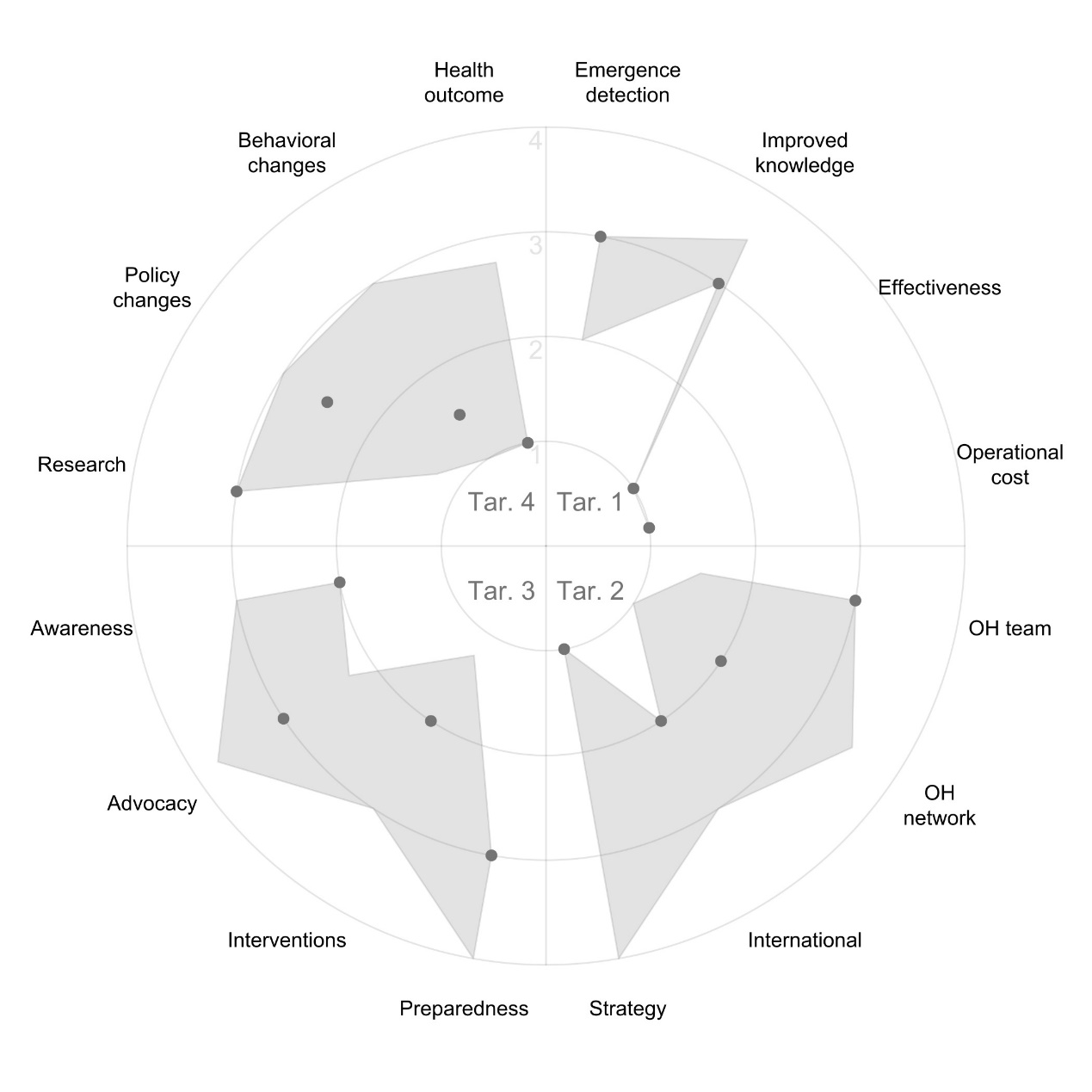

Figure S2_3: OH-EpiCap Dimension three (impact) visualised at the indicator level. Plotted points indicate the median score with interquartile range indicated by the shaded polygons across four targets: target 1: technical outputs; target 2: collaborative added-value; target 3: immediate and intermediate outcomes; target 4: ultimate outcomes.
