## Supplementary File S1 for "Implementation of One Health surveillance systems: opportunities and challenges - Lessons learned from the OH-EpiCap application"

**Supplementary table 1:** List of the institutes that participated in the OH-EpiCap evaluation of their specific surveillance system

| **Country** | **Hazard** | **Institutes Participated in the evaluation** | **OH-Epicap evaluation** |
| --- | --- | --- | --- |
| Denmark | Psittacosis | **Animal health:** Danish Veterinary and Food Administration (Fødevarestyrelsen)  **Public health:** Statens Serum Institut | Workshop (seven surveillance representatives) |
| Finland | *Listeria* | **Food safety:** Finnish Food Safety Authority (Ruokavirasto)  **Public health:** Finnish Institute for Health and Welfare (THL) | Workshop (five surveillance representatives) |
| France | AMR | **Animal health / Food safety:** National Agency for Food, Environmental and Occupational Health and Safety (ANSES)  **Public health:** Directorate General for Food (DGAL) and National Public Health Agency (SpF) | Questionnaire filled sequentially by three surveillance representatives |
| France | *Salmonella* | **Food safety / Public health:** National Agency for Food, Environmental and Occupational Health and Safety (ANSES) | Questionnaire filled by one surveillance representative |
| Germany | *Salmonella* | **Animal health:** Friedrich-Loeffler-Institut (FLI)  **Food safety:** Federal Institute for Risk Assessment (BfR)  **Public health:** Robert Koch Institute (RKI) | Workshop (ten surveillance representatives) |
| Norway | *Campylobacter* | **Animal health / Food safety:** Norwegian Veterinary Institute (NVI)  **Food safety:** Norwegian Food Safety Authority | Questionnaire filled sequentially by two surveillance representatives |
| Norway | *Listeria* | **Animal health:** Norwegian Veterinary Institute (NVI)  **Food safety:** Norwegian Food Safety Authority | Questionnaire filled sequentially by two surveillance representatives |
| Portugal | AMR | **Public health:** Directorate General for Health (DGS) and National Institute of Health Doctor Ricardo Jorge (INSA)  **Animal health:** National Institute of Agricultural and Veterinary Investigação (INIAV)  **Environment:** Portuguese Environment Agency (APA) | Questionnaire filled sequentially by four surveillance representatives |
| Sweden | *Campylobacter* | **Animal health:** National Veterinary Institute (SVA)  **Food safety:** Swedish National Food Agency (SLV), Swedish Board of Agriculture (Jordbruksverket)  **Public health:** Public Health Agency of Sweden (Folkhalsomyndigheten) | Workshop (five surveillance representatives) |
| The Netherlands | *Listeria* | **Public Health:** National Institute for Public Health and the Environment (RIVM) | Questionnaire filled by two surveillance representatives |
| The Netherlands | *Salmonella* | **Public Health:** National Institute for Public Health and the Environment (RIVM) | Questionnaire filled by two surveillance representatives |
